# Pre-Operative Single 150 Mg Dose of Pregabalin for Postoperative Pain Management in Laparoscopic Cholecystectomy: A Systematic Review and Meta-Analysis

**DOI:** 10.64898/2026.07.11.26357848

**Authors:** Ghisaram Dewasi, Parikshit Nagda, Sakshi Jain, Shivam Soni

## Abstract

Effective postoperative pain control is essential following laparoscopic cholecystectomy, yet the analgesic value of a standardised 150 mg preoperative dose of pregabalin has not been clearly established. This systematic review and meta-analysis synthesised evidence from seven randomised controlled trials published between 2008 and 2025 to evaluate the efficacy and safety of pregabalin when administered before surgery. Four trials reported 24-hour postoperative pain scores, and pooled analysis demonstrated that pregabalin significantly reduced pain compared with control (SMD = –0.80; 95% CI: –1.42 to –0.18; *p* = .01), although heterogeneity was high (I² = 81%). Pregabalin also produced notable reductions in opioid consumption, including fentanyl (SMD = –1.24; *p* = .002) and tramadol (SMD = –4.21; *p* = .002), again with considerable variability across studies. Sedation was slightly increased but did not reach statistical significance, and there were no significant differences in postoperative nausea, vomiting, or headache. Sensitivity analyses supported the stability of these findings. Overall, the results indicate that a single 150 mg preoperative dose of pregabalin meaningfully reduces postoperative pain and opioid requirements following laparoscopic cholecystectomy while maintaining an acceptable safety profile, supporting its use as part of a multimodal analgesic strategy.

## 1. Introduction

Postoperative pain remains an important clinical challenge following laparoscopic cholecystectomy (LC), with up to 80% of patients experiencing moderate-to-severe acute pain within 24 hours. This not only compromises recovery but also increases hospital stays and healthcare costs. While opioids are conventionally an important part of postoperative analgesia, their use is limited by adverse effects, including respiratory depression, nausea, and the risk of dependence. Consequently, multimodal analgesia integrating non-opioid adjuncts has emerged as a gold standard to enhance pain control while minimising opioid-related complications (1–3)

An ideal pre-emptive analgesic should exhibit anxiolytic, sedative, and opioid-sparing properties while reducing surgical stress responses. Pregabalin, a gabapentinoid, has gained attention for its potential in early postoperative pain management. Its mechanism suppresses the release of excitatory neurotransmitters (binds to the α δ subunit of voltage-gated calcium channels in the central nervous system), thereby inhibiting hyperalgesia and central sensitisation. Compared to gabapentin, pregabalin offers superior pharmacokinetics: linear absorption, higher bioavailability, and fewer drug interactions (4–7)

Clinical studies report that a single pre-emptive dose of 150mg of pregabalin before LC reduces early postoperative pain scores, decreases opioid consumption by 30–40%, and improves sedation quality without significant adverse effects. For instance, a randomised controlled trial (RCT) by Jokela et al. (2008) demonstrated that 150 mg pregabalin preoperatively lowered 24-hour morphine use by 38% and reduced dynamic pain scores. However, conflicting evidence exists for low doses. Studies like Peng et al. conducted a randomised controlled trial using low-dose pregabalin (50 mg and 75 mg) (8) found only modest early pain relief, with no significant reduction in overall analgesic use, reflecting the limited value of low-dose pregabalin as a stand-alone pre-emptive analgesic (2,7,9,10)

Given the clinical importance of identifying an optimal pregabalin regimen, we conducted a systematic review and meta-analysis to evaluate the efficacy and safety of a single preoperative 150 mg oral dose of pregabalin in patients undergoing laparoscopic cholecystectomy.

## 2. Materials and Methods

This systematic review and meta-analysis were performed following the Preferred Reporting Items for Systematic Reviews and Meta-Analyses (PRISMA) guidelines and the Cochrane Handbook for Systematic Reviews of Interventions. Ethical approval and patient consent were not required, as the analysis was based on previously published studies (11, 12).

### Literature search

From 2008 to May 2025, Google Scholar, Web of Science, PubMed, and the Cochrane Library were searched using specific keywords (pregabalin, laparoscopic cholecystectomy, postoperative pain), and relevant articles were extracted from these databases. Identified articles were screened using titles, abstracts, and keywords. The meta-analysis collected data from the published articles; hence, no ethical approval was needed.

### Inclusion Criteria and Study Selection

We determined the inclusion criteria following the PICOS principle (13).

Population: Patients undergoing laparoscopic cholecystectomy

Intervention: preoperative 150mg single oral dose of pregabalin was used as an adjunct to multimodal anaesthetics

Comparison: Placebo

Outcome: Pain scores at 24 hours, total fentanyl consumption, Total tramadol consumption and related side effects like nausea, vomiting and headache.

Study Design: Randomised Controlled Trials

Two individual reviewers screened the articles after the duplicate removal. Selected articles are matched to the inclusion criteria, and fail to match the inclusion criteria are removed, and any disagreements about the inclusion or exclusion of studies were resolved by discussion with an expert.

### Data Extraction and Quality assessment in individual Studies

A Microsoft Excel spreadsheet is used to collect the data from the selected RCTs. The data was extracted as follows: First author and publication year, sample size of pregabalin and placebo groups, timing of the dose and demographic data of the patients included in the studies. Outcomes such as VAS score (10cm) at 2h and 24 hours (Mean±SD), total fentanyl consumption, tramadol consumption, first time to rescue analgesic, postoperative nausea, vomiting, headache and sedation (Ramsay sedation scale) are extracted from the studies. These outcomes are classified as primary (VAS score) and secondary outcomes (other data).

To assess the methodological quality of each RCTs, including Jadad scale is used. This scale has three evaluation elements: randomisation (0-2 points), blinding (0-2 points), dropouts and withdrawals (0-1 point) (Jadad scale reference). One point would be allocated to each element if they are mentioned in the articles, and the remaining 2 points are given if the randomisation method and blinding are explained in detail. If the randomisation and/ or blinding were inappropriate, or dropouts are not mentioned, then one point is deducted. The score varies from 0-5 points. A Jadad score≤2 was considered to be of low quality. If the Jadad score≥3, the study was thought to be of high quality (14).

### Outcome measures and statistical analysis

Outcome measures (VAS score at 24 hours, total fentanyl, tramadol consumption) were expressed in the Standard mean difference (SMD) with 95% confidence interval. Dichotomous outcomes (Nausea, vomiting and headache) were expressed as risk ratio (RR) with 95% confidence interval. The statistical significance is set at P<0.05. Variables in the meta-analysis were calculated using Review Manager 5.4.1 (The Cochrane Collaboration, Software Update, Oxford, UK). We used a random effect model to summarise the outcome of our study. Heterogeneity is also assessed by the Cochran Q statistics and quantified with I^2^ statistics. An I^2^ value >50% indicates significant heterogeneity. A leave-one-out sensitivity analysis was conducted, where each study was sequentially removed to determine its influence on the overall effect size. A funnel plot was not generated to assess the publication bias, as fewer than 10 studies were included (15).

## 3. Results

### 3.1. Literature search and Quality assessment

In the initial search, a total of (Fig 1.) 495 studies were identified from the electronic database (PubMed: 102, Cochrane: 187, Google Scholar: 108, Web of Science: 98). All papers were input into the Rayyan web application for the removal of duplicate papers. A total of 285 papers were reviewed, and 266 papers were removed according to the inclusion and exclusion criteria by reading abstracts and titles. Ultimately, 7 randomised controlled trials with 450 patients (pregabalin= 225, Placebo= 225) (16–22). Only RCTs with 150mg pregabalin preoperatively included. General characteristics of the included studies can be seen in Table 1. Among the seven RCTs, four reported pain score (16–19), four reported fentanyl consumption (16,18,20,21), two reported tramadol consumption (17,19), three reported Ramsay sedation scale (16,17,21), four studies reported postoperative nausea and vomiting (16–18,21), and two studies reported headache (16,18). Jadad scores of included studies vary from 3-5, which shows that the studies were high-quality studies.

**Fig. 1.**
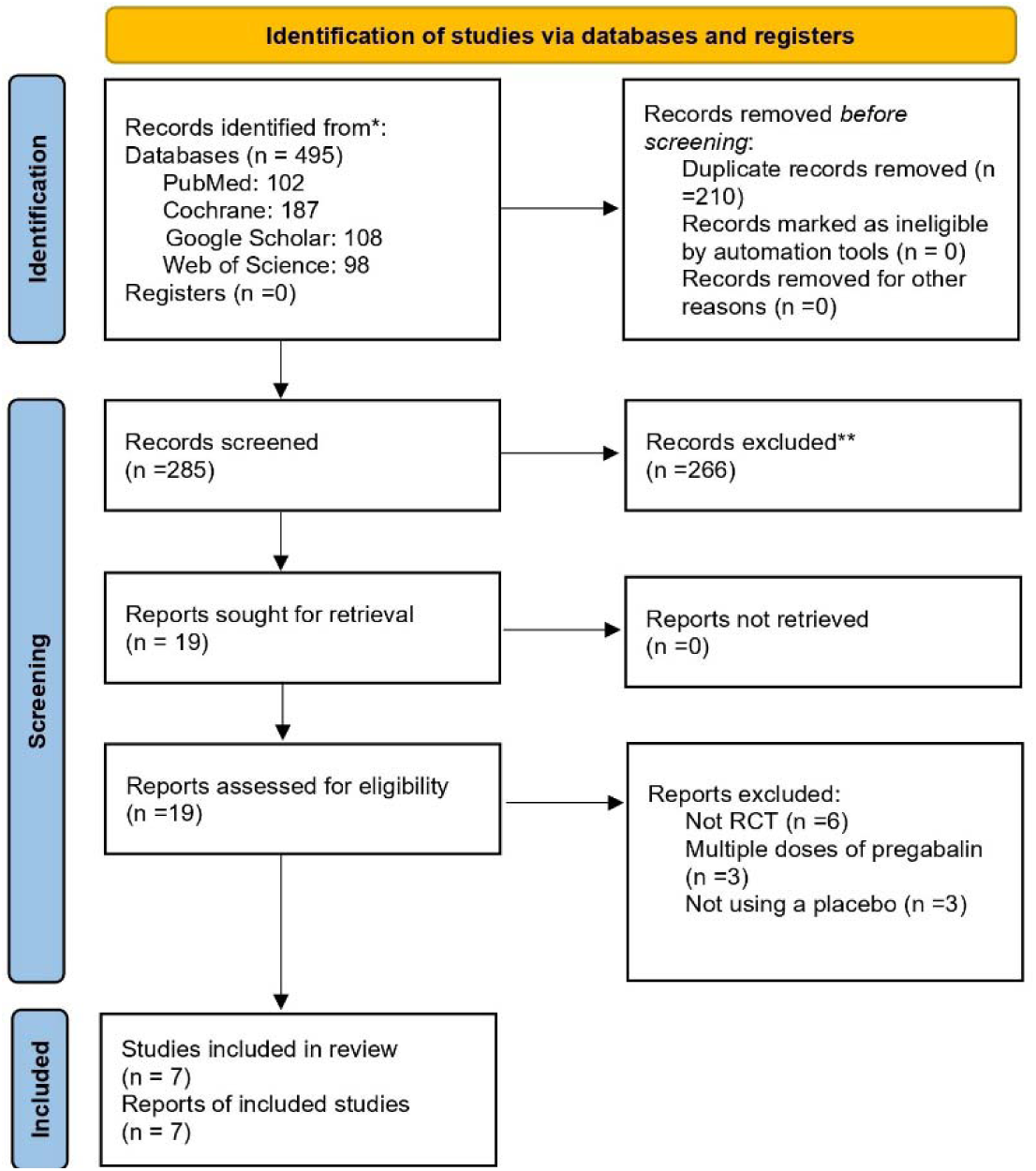
Flow diagram of the study searching and selection process

Table 1: General Characteristics of Studies:

### 3.2. Primary Outcomes

The primary outcome of this meta-analysis was to assess the overall effect of pregabalin compared to a placebo on the primary measure (VAS scores).

#### 3.2.1. VAS scores 24 hours

Among the seven RCTs, four studies reported the VAS score at multiple time intervals. we collected the data for 24 hours to assess the 24-hour analgesic effect of a single dose of the pregabalin.

Data is analysed with a random effects model. The pooled estimate of the four included randomised controlled trials suggested that preoperative administration of a single 150mg dose of pregabalin can decrease the VAS score at 24 hours. The pooled standardised mean difference (SMD) using a random-effects model was -0.80 with a 95% confidence interval (CI) of [-1.42, -0.18], indicating a statistically significant effect in favour of pregabalin (*Z* = 2.55, *p* = .01) (Fig. 2). Heterogeneity among the included studies was substantial (I² = 81%, *p* = .001), suggesting variability in effect sizes beyond chance.

**Fig. 2.**
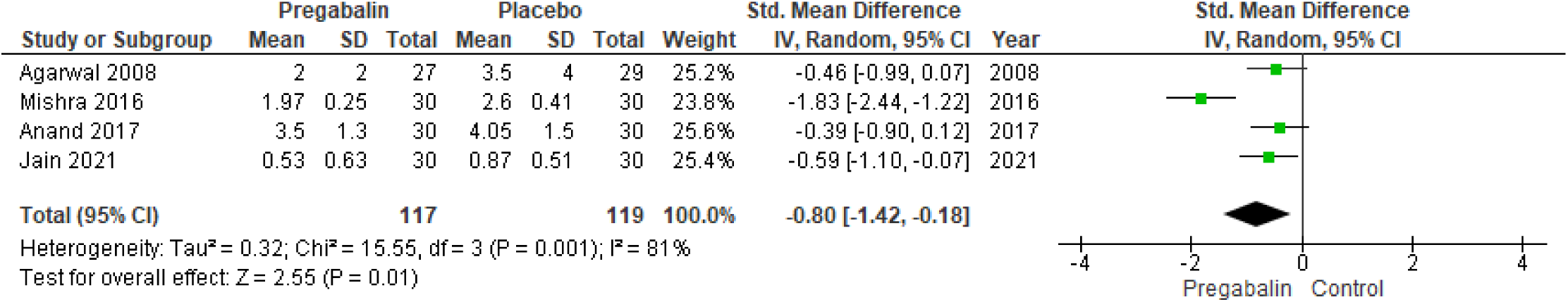
Forest plot for the meta-analysis of pain score 24 hours.

#### 3.2.1. Sensitivity test

To evaluate the reliability of the results, a leave-one-out sensitivity analysis was conducted (Fig.3), where each study was sequentially removed to determine its influence on the overall effect size. The sensitivity analysis results revealed that the pooled effect size remained relatively stable, with most studies still producing statistically significant effects after exclusion. The exception was the Jain (2021) (18) study, where the p-value became marginally significant (0.050). This study’s exclusion had a minor impact on the pooled effect, suggesting that its influence on the overall result is weaker compared to the other studies.

**Fig. 3.**
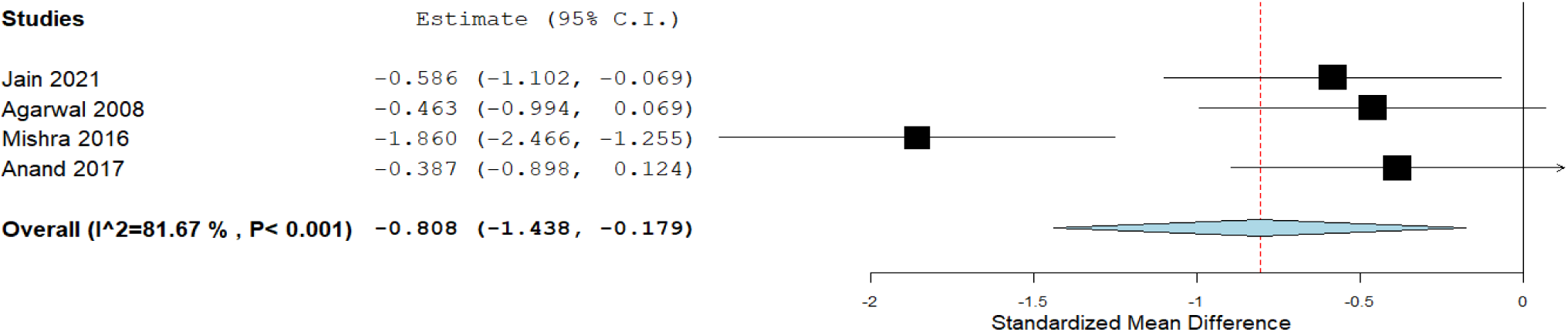
Forest plot for a leave-one-out sensitivity analysis

Overall, the findings from the leave-one-out sensitivity analysis support the strength of the pooled effect size, demonstrating that the conclusions drawn from the meta-analysis are not heavily dependent on any single study. The results indicate that pregabalin has a statistically significant and consistent effect in reducing VAS scores across studies.

### 3.3 Secondary Outcomes

Meta-analysis revealed that pregabalin compared to placebo significantly reduced both fentanyl (SMD = –1.24, 95% CI [–2.03, –0.44], *p* = .002, I² = 87%; Figure.4) and tramadol consumption (SMD = –4.21, 95% CI [–6.82, –1.60], *p* = .002, I² = 93%; Figure.5) compared to placebo, though both showed high heterogeneity. Ramsay sedation scores showed a non-significant increase with pregabalin (SMD = 0.36, 95% CI [–0.06, 0.78], *p* = .09, I² = 49%: Figure 6).

**Fig. 4.**
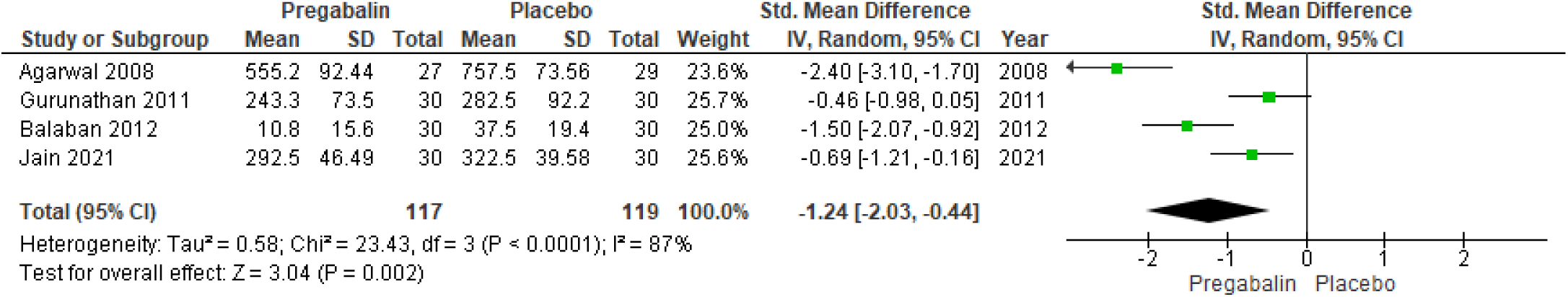
Forest plot of Fentanyl consumption in Pregabalin and Placebo groups

**Fig. 5.**
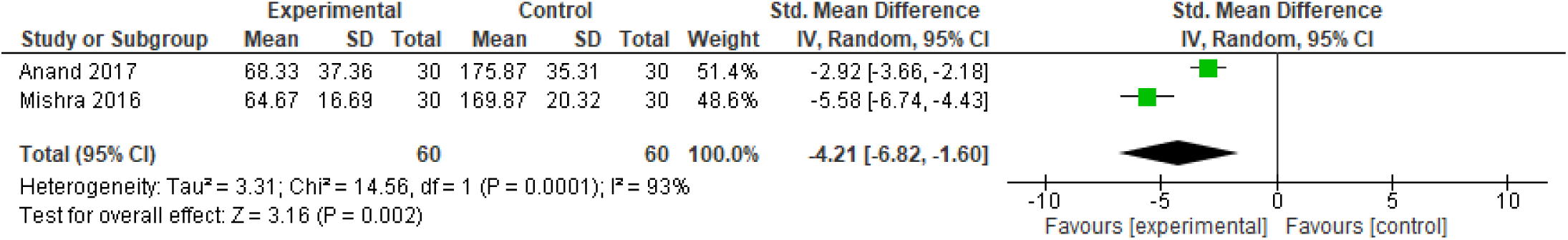
Forest plot of Tramadol Consumption in Pregabalin and Placebo groups

**Fig. 6.**
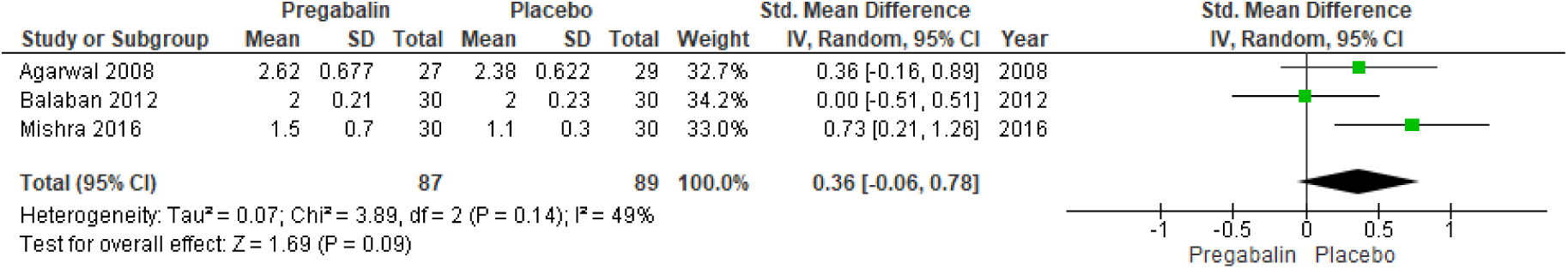
Forest plot of Ramsay sedation score between pregabalin and placebo groups

**Fig. 7.**
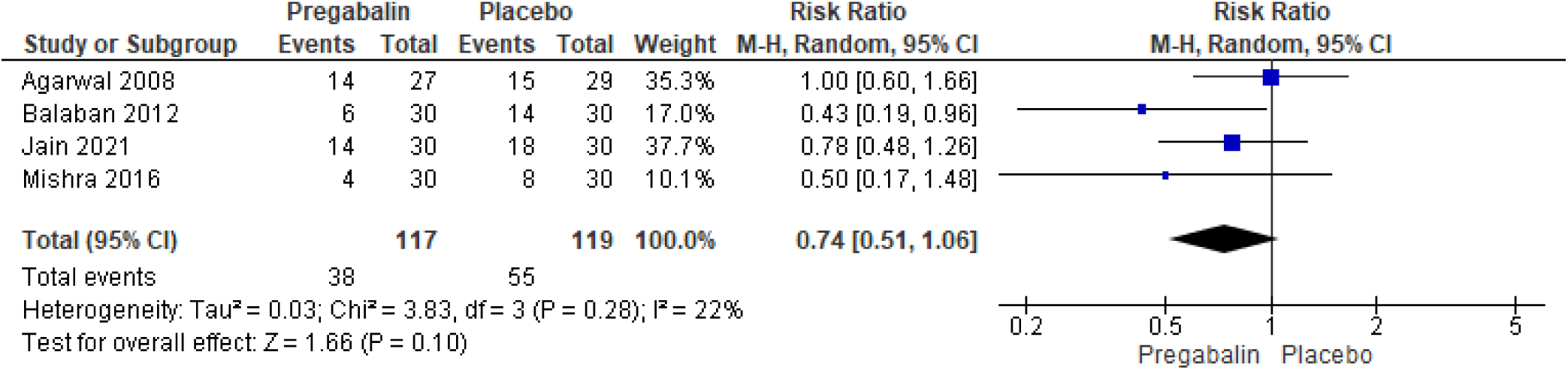
Forest plot of post-operative nausea and vomiting in the pregabalin and placebo groups

**Fig. 8.**
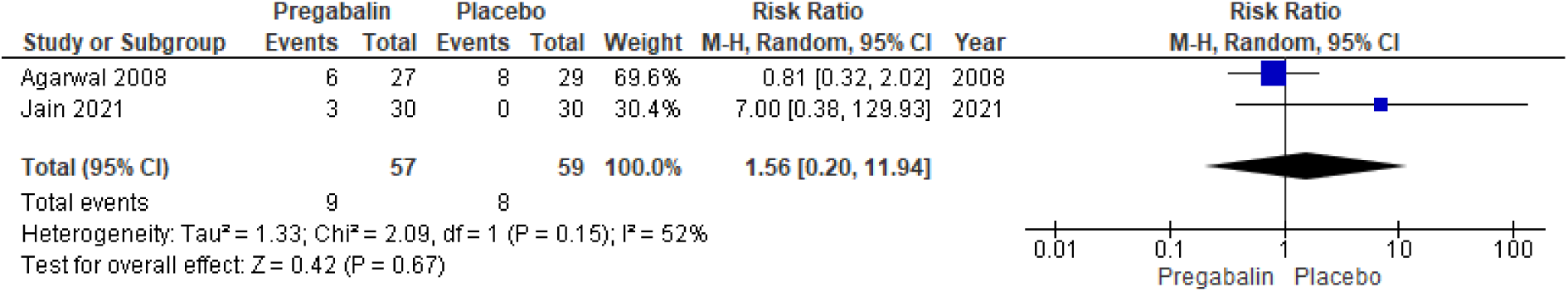
Forest plot of Headache in pregabalin and placebo groups

For adverse effects, pregabalin showed a non-significant reduction in the risk of postoperative nausea and vomiting (PONV) (RR = 0.74, 95% CI [0.51, 1.06], *p* = .10, I² = 22%; Figure.7) and a non-significant increase in headache risk (RR = 1.56, 95% CI [0.20, 11.94], *p* = .67, I² = 52% Figure.8).

## 4. Discussion

This meta-analysis evaluated the efficacy and safety of a single preoperative 150 mg dose of pregabalin in patients undergoing laparoscopic cholecystectomy (LC). The pooled data from high-quality randomized controlled trials (RCTs; Jadad score>3) revealed that pregabalin significantly reduced postoperative pain at 24 hours, as evidenced by a moderate effect size in VAS scores (SMD = –0.80, 95% CI [–1.42, –0.18], *p* = .01) potentially reducing the need for rescue analgesics. These findings align with the pharmacological rationale of pregabalin, which acts centrally by modulating calcium channels and inhibiting nociceptive neurotransmitter release, thereby attenuating central sensitisation and hyperalgesia (23–25).

Importantly, the opioid-sparing effects of pregabalin were evident across multiple studies. Our analysis showed a statistically significant reduction in both fentanyl (SMD = –1.24, 95% CI [–2.03, –0.44]) and tramadol consumption (SMD = –4.21, 95% CI [–6.82, –1.60]) in the pregabalin group compared to placebo. These outcomes are consistent with earlier reports demonstrating reduced opioid requirements and improved recovery profiles when pregabalin is used as an adjunct to multimodal analgesia (16,17,21). Given the global imperative to reduce opioid-related complications, such findings have strong clinical relevance.

Regarding sedation, although Ramsay sedation scores increased slightly in the pregabalin group (SMD = 0.36), the difference was not statistically significant (*p* = .09), suggesting that 150 mg pregabalin provides mild anxiolysis without inducing excessive sedation. This finding is clinically desirable as excessive sedation may delay ambulation and recovery. Furthermore, pregabalin demonstrated a non-significant trend toward reduced postoperative nausea and vomiting (RR = 0.74, *p* = .10), and while headache incidence was slightly higher (RR = 1.56), this was not statistically significant (*p* = .67). These outcomes support pregabalin’s safety at the 150 mg dose level, aligning with earlier evidence reporting low incidence of serious adverse events at this dosage (8,9). Additionally, in most included studies, pregabalin was administered 1 to 2 hours before the surgery, which appears to be an optimal timing window to achieve effective plasma concentrations while minimising sedative side effects.

From a mechanistic standpoint, the analgesic efficacy of 150 mg pregabalin is attributed to its optimal balance between bioavailability and central nervous system penetration, without the diminishing returns or increased side effects seen at higher doses (≥300 mg) (24–26). On the other hand, lower doses (50–75 mg) have demonstrated limited efficacy in previous trials, failing to reduce pain or opioid use consistently (17). Thus, our findings suggest that the integration of pregabalin 150 mg preoperatively into a routine multimodal analgesia protocol enhances the recovery and reduces opioid reliance. These results align with enhanced recovery after surgery principles by reducing the opioid use, faster recovery and improved patient comfort.

Despite the encouraging findings, certain limitations must be acknowledged. The heterogeneity observed in primary (I² = 81%) and secondary outcomes (e.g., tramadol I² = 93%) reflects clinical and methodological variability, such as differences in surgical technique, anaesthesia protocols, or baseline patient characteristics. Nonetheless, sensitivity analysis confirmed the robustness of the pooled estimates, with no single study disproportionately influencing the results. Moreover, the absence of a publication bias assessment may limit generalizability due to the inclusion of fewer than 10 studies (27,28).

## 5. Conclusion

A single preoperative oral dose of 150 mg pregabalin appears to be an effective and well-tolerated adjunct for perioperative analgesia in patients undergoing laparoscopic cholecystectomy. It significantly reduces postoperative pain intensity and opioid consumption and post-operative nausea & vomiting without increasing the risk of sedation, or other common adverse effects. These findings support the integration of pregabalin into multimodal analgesia protocols, particularly within enhanced recovery frameworks. Given its favourable efficacy-safety profile and ease of administration, pregabalin may serve as a practical strategy to optimise postoperative outcomes in minimally invasive general surgery. Further prospective trials are warranted to validate these findings across broader surgical populations and inform evidence-based clinical guidelines.

## Declaration

### Ethical Considerations

Not applicable

## Author contribution

All authors are equal to this work.

## Availability of data and material

Not applicable.

## Data Availability

Data is available upon the reasonable request to corresponding author.

## Acknowledgements

Not applicable.

The Open Science Framework (OSF) registration: <u>osf.io/m89d5</u>

## List of the abbreviations

Abbreviation: Full Form
LC: Laparoscopic Cholecystectomy
RCT: Randomized Controlled Trial
VAS: Visual Analogue Scale
PONV: Postoperative Nausea and Vomiting
SMD: Standardized Mean Difference
RR: Risk Ratio
CI: Confidence Interval
PRISMA: Preferred Reporting Items for Systematic Reviews and Meta-Analyses OSF Open Science Framework
CNS: Central Nervous System
α δ: Alpha-2-delta subunit
Jadad: Oxford Quality Scoring System
I²: I-squared (Statistical measure of heterogeneity)
NCBI: National Center for Biotechnology Information
UK: United Kingdom

## References

1. Najam F, Jafri N, Khan MN, Daraz U. Reduction of Acute Postoperative Pain With Pre-Emptive Pregabalin Following Laparoscopic Cholecystectomy. Cureus. 2022 Aug;14(8):e27783.

2. Hazarika R, Talukdar FA, Bharali H, Bhattacharjee R. PREEMPTIVE SINGLE-DOSE PREGABALIN IN MODULATION OF POSTOPERATIVE PAIN AND OPIOID REQUIREMENT AFTER LAPAROSCOPIC CHOLECYSTECTOMY- A RANDOMIZED CLINICAL STUDY. J Evid Based Med Healthc. 2018 Jan 22;5(4):319–25.

3. Zhang D, You G, Yao X. Influence of pregabalin on post-operative pain after laparoscopic cholecystectomy: A meta-analysis of randomised controlled trials. J Minimal Access Surg. 2020;16(2):99–105.

4. Zhang J, Ho KY, Wang Y. Efficacy of pregabalin in acute postoperative pain: a meta-analysis. Br J Anaesth. 2011 Apr;106(4):454–62.

5. (PDF) Effects of Pregabalin versus Gabapentin on their Opioid Sparing Effects among Patients Undergoing Laproscopic Cholecystectomy: A Randomised Controlled Study [Internet]. [cited 2025 Jun 24]. Available from: https://www.researchgate.net/publication/363277580_Effects_of_Pregabalin_versus_Gabapentin_on_their_Opioid_Sparing_Effects_among_Patients_Undergoing_Laproscopic_Cholecystectomy_A_Randomised_Controlled_Study

6. Pregabalin - StatPearls - NCBI Bookshelf [Internet]. [cited 2025 Jun 24]. Available from: https://www.ncbi.nlm.nih.gov/books/NBK470341/

7. Bekawi MS, El Wakeel LM, Al Taher WMA, Mageed WMA. Clinical study evaluating pregabalin efficacy and tolerability for pain management in patients undergoing laparoscopic cholecystectomy. Clin J Pain. 2014 Nov;30(11):944–52.

8. Peng PWH, Li C, Farcas E, Haley A, Wong W, Bender J, et al. Use of low-dose pregabalin in patients undergoing laparoscopic cholecystectomy. Br J Anaesth. 2010 Aug;105(2):155–61.

9. Jokela R, Ahonen J, Tallgren M, Haanpää M, Korttila K. A randomized controlled trial of perioperative administration of pregabalin for pain after laparoscopic hysterectomy. Pain. 2008 Jan 1;134(1):106–12.

10. Zhang Y, Wang Y, Zhang X. Effect of pre-emptive pregabalin on pain management in patients undergoing laparoscopic cholecystectomy: A systematic review and meta-analysis. Int J Surg Lond Engl. 2017 Aug;44:122–7.

11. Preferred reporting items for systematic reviews and meta-analyses: the PRISMA statement | The BMJ [Internet]. [cited 2025 Jun 25]. Available from: https://www.bmj.com/content/339/bmj.b2535

12. Home - The Peptide Report [Internet]. [cited 2025 Jun 25]. Available from: https://www.thepeptidereport.com/

13. Eldawlatly A, Alshehri H, Alqahtani A, Ahmad A, Al-Dammas F, Marzouk A. Appearance of Population, Intervention, Comparison, and Outcome as research question in the title of articles of three different anesthesia journals: A pilot study. Saudi J Anaesth. 2018;12(2):283–6.

14. Reported Methodologic Quality and Discrepancies between Large and Small Randomized Trials in Meta-Analyses | Annals of Internal Medicine [Internet]. [cited 2025 Jun 24]. Available from: https://www.acpjournals.org/doi/10.7326/0003-4819-135-11-200112040-00010

15. Nair AS. Publication bias - Importance of studies with negative results! Indian J Anaesth. 2019 Jun;63(6):505–7.

16. Agarwal A, Gautam S, Gupta D, Agarwal S, Singh P, Singh U. Evaluation of a single preoperative dose of pregabalin for attenuation of postoperative pain after laparoscopic cholecystectomy. 2008;101(5):700 704.

17. Mishra R, Tripathi M, Chandola HC. Comparative clinical study of gabapentin and pregabalin for postoperative analgesia in laparoscopic cholecystectomy. Anesth Essays Res. 2016;10(2):201–6.

18. Singh T, Kathuria S, Jain R, Sood D, Gupta S. Premedication with pregabalin 150mg versus 300mg for postoperative pain relief after laparoscopic cholecystectomy. J Anaesthesiol Clin Pharmacol. 2020;36(4):518–23.

19. Archives | Anaesthesia, Pain & Intensive Care [Internet]. [cited 2025 Jun 24]. Available from: https://www.apicareonline.com/index.php/APIC/issue/archive

20. Gurunathan U, Rapchuk IL, King G, Barnett AG, Fraser JF. The effect of pregabalin and celecoxib on the analgesic requirements after laparoscopic cholecystectomy: a randomized controlled trial. J Anesth. 2016 Feb;30(1):64–71.

21. Balaban F, Yağar S, Özgök A, Koç M, Güllapoğlu H. A randomized, placebo-controlled study of pregabalin for postoperative pain intensity after laparoscopic cholecystectomy. 2012;24(3):175–178.

22. Gupta K, Sharma D, Gupta PK. Oral premedication with pregabalin or clonidine for hemodynamic stability during laryngoscopy and laparoscopic cholecystectomy: A comparative evaluation. Saudi J Anaesth. 2011;5(2):179–84.

23. Gee NS, Brown JP, Dissanayake VU, Offord J, Thurlow R, Woodruff GN. The novel anticonvulsant drug, gabapentin (Neurontin), binds to the alpha2delta subunit of a calcium channel. J Biol Chem. 1996 Mar 8;271(10):5768–76.

24. Bockbrader HN, Wesche D, Miller R, Chapel S, Janiczek N, Burger P. A comparison of the pharmacokinetics and pharmacodynamics of pregabalin and gabapentin. Clin Pharmacokinet. 2010 Oct;49(10):661–9.

25. Imani F, Rahimzadeh P. Gabapentinoids: Gabapentin and Pregabalin for Postoperative Pain Management. Anesthesiol Pain Med. 2012;2(2):52–3.

26. Dewasi, G., Nagda, P., Bahl, G. et al. Myostatin-driven muscle hypertrophy: a double-edged sword in muscle physiology. J Rare Dis 4, 29 (2025). 10.1007/s44162-025-00086-x

27. Tanwar CS, Jain V, Nagda P, Ramani K, Bahl G, Dewasi G, et al. A Rare Presentation of chronic lower back ache radiating to lower limb in a patient with Ankylosing Spondylitis with Andersson Lesion managed by Methotrexate: A Case Report [Internet]. In Review; 2025 [cited 2026 Aug 8]. Available from: https://www.researchsquare.com/article/rs-6459382/v1 doi:10.21203/rs.3.rs-6459382/v1

28. Dewasi G, Vaghela JS, Saraf M, Jain S. Evaluation of preemptive single dose of pregabalin for postoperative pain control after laparoscopic cholecystectomy: a prospective comparative study from western India. Dolor® Investig Clínica Ter. 2026. doi:10.24875/DOL.M25000032

